# Comparison of plasma soluble and extracellular vesicles-associated biomarkers in Alzheimer’s Disease patients and cognitively normal individuals

**DOI:** 10.1101/2024.02.26.24303378

**Authors:** Emilien Boyer, Louise Deltenre, Marion Dourte, Lise Colmant, Esther Paître, Kristel Sleegers, Nuria Suelves, Bernard Hanseeuw, Pascal Kienlen-Campard

## Abstract

**INTRODUCTION:** Amyloid-β (Aβ) and tau are brain hallmarks of Alzheimer’s disease (AD) also present in blood as soluble biomarkers or encapsulated in extracellular vesicles (EVs). Our goal was to assess how soluble plasma biomarkers of AD pathology correlate with number and content of EVs.

**METHODS:** Single-molecule enzyme-linked assays were used to quantify Aβ42/40 and tau in plasma samples and neurally-derived EVs (NDEVs) from a cohort of *APOE* ε4– and *APOE* ε4+ cognitively normal individuals (CN) and AD patients.

**RESULTS:** Soluble plasma Aβ42/40 ratio is decreased in AD patients compared to CN individuals. The amount and content (Aβ40, Aβ42, tau) of plasma NDEVs were similar between groups. Quantity of soluble biomarkers were negatively correlated to NDEVs number only in CN individuals.

**DISCUSSION:** Soluble Aβ42/40 ratio is the most robust AD plasma biomarker. Analysis of NDEVs and their content pointed toward peculiar mechanisms of Aβ release in AD.

**Institutional Review Board Statement:** The study was conducted in accordance with the Declaration of Helsinki and approved by the Institutional Ethics Committee of Clinics Saint-Luc University Hospital, 1200 Brussels, Belgium (UCL-2022-473; UCL-2016-121; UCL-2018-119).

**HIGHLIGHTS:**

- The number of neurally-derived extracellular vesicles (NDEVs) in plasma is not a stand-alone biomarker for Alzheimer’s disease (AD).
- Plasma levels of Aβ42, Aβ40 and total-tau are strongly negatively correlated with NDEVs concentration in cognitively normal (CN) individuals.
- In AD patients, this correlation is lost, highlighting a shift in the mechanism underpinning the production and the release of these biomarkers in pathological conditions.
- The soluble plasma amyloid-β (Aβ) 42/40 ratio is the most robust biomarker to discriminate between AD patients and CN individuals, as it normalizes for the number of NDEVs.

**RESEARCH IN CONTEXT:**

1. **Systematic review:** The authors reviewed PubMed for plasma biomarkers and neurally-derived extracellular vesicles (NDEVs) in AD. While many studies focus on soluble plasma AD biomarkers like Aβ42/40 ratio or tau protein variants, less explore NDEVs. This study investigates NDEVs as potential AD biomarkers and their correlation with soluble plasma biomarkers.
2. **Interpretation:** Our study confirms decreased soluble Aβ42 and increased soluble total-tau in AD patients, with soluble Aβ40 indicating elevated Aβ production in AD versus CN individuals. The Aβ42/40 ratio is a robust AD biomarker. CN individuals with AD risk (*APOE* ε4+) show decreased ratios without symptoms. Plasma NDEVs remain consistent across ages and between AD and CN individuals, but correlations with soluble plasma biomarkers suggest altered Aβ processes in AD.
3. **Future directions:** Further research on independent cohorts can confirm our findings and assess whether plasma Aβ and tau need correction by NDEVs for better AD risk identification in CN populations.

## 1. Introduction

The prevalence of age-related neurodegenerative diseases is expected to increase globally in the coming decades, attributed to improved overall health conditions and increased life expectancy in many countries. Currently, around 50 million individuals are affected by dementia, a number projected to reach 130 million by 2050[1]. Alzheimer’s disease (AD) is the primary cause of senile dementia. Finding reliable, easily accessible, and cost-effective biomarkers is a major challenge for the accurate and early diagnosis of AD. Diagnostic techniques based on neuroimaging and fluidic biomarkers have made great strides over the last two decades. New tracers like the ^18^F-Flortaucipir have been developed to accurately image tau pathology in the brain and are approved by the FDA[2]. However, the cost and limited accessibility of PET imaging to clinical research centers prevent, for instance, large-scale screening of at-risk populations.

Regarding fluidic biomarkers, the analysis of amyloid-β peptide (Aβ42), tau, and phospho-tau (p-tau) in cerebrospinal fluid (CSF) has long been the gold standard for AD diagnosis[3]. These biomarkers are also key actors of AD pathological process, and prime targets for the development of disease-modifying therapies. Monoclonal antibodies targeting aggregated Aβ, such as aducanumab and lecanemab, gained FDA approval in 2021 and 2023[4, 5]. These emerging disease-modifying therapies might greatly benefit from screening techniques that are more accessible, less invasive than lumbar puncture and that would accurately inform about disease progression While blood-based biomarkers hold promise, none is currently fully validated or clinically used for AD. The development of blood biomarkers faces certain challenges. For example, the low plasma concentrations of central nervous system (CNS)-derived biomarkers require a robust and reliable technique for their detection that can be implemented for routine use. The development of ultrasensitive single molecule detection arrays[6] has set new standards for biomarker detection in plasma or serum, fueling the interest for the easily-accessible blood samples. Still, regardless of their sensitivity, these quantification techniques detect only soluble/monomeric forms of Aβ and tau, leaving aside other forms (e.g. aggregated or encapsulated in biological vesicles) that may be relevant for understanding the pathological processes. Other parameters can additionally act as confounding factors in biomarker quantification. Sex seems to influence plasma amyloid, as evidenced by a lower Aβ42/40 ratio in males[7]. Age is also a factor, as older cognitively normal (CN) individuals show lower levels of soluble Aβ42 and Aβ42/40 ratio[7–9]. *APOE* status is associated with amyloid levels, with significantly lower Aβ42 levels in *APOE* ε4 carriers in CN individuals[7, 9]. Additionally, factors like body mass index, recently linked to plasma concentrations of Aβ42 and Aβ40, should be considered as potential influencers of Aβ levels in blood[10].

Recent investigations indicate that extracellular vesicles (EVs) are highly transmissible and play critical roles in the propagation of tau pathology [11]. Although EVs’ precise role in AD pathogenesis remain elusive, they may facilitate the spreading of pathological factors like Aβ and tau seeds between cells[12, 13]. Other studies showed that EVs can be loaded with neurotoxic Aβ forms[14]. The processing of the Amyloid Precursor Protein (APP) occurs in late endosomal pathways, involved EV formation, which could be a potential source of Aβ release in the extracellular space[15, 16]. Neurally-derived EVs (NDEVs), detectable in plasma, can cross the blood-brain barrier. NDEVs appear thus as a promising tool to investigate brain changes during disease progression, with their content potentially reflecting AD pathogenesis more accurately than soluble blood biomarkers or providing complementary information[12].

Our aim in this study was to measure with high accuracy and sensitivity Aβ peptides, tau and EVs in plasma samples and evaluate how they correlate to the clinical status of cognitively normal or AD individuals. Furthermore, we aimed at determining whether the concentration of NDEVs present in the plasma can (i) reflect AD progression, (ii) modulate the levels of soluble biomarkers, and can be used as a potential diagnosis tool and as an indication of possible changes in the mode of Aβ production associated to AD pathogeny.

## 2. Methods

### 2.1. Participants and study settings

Participants were recruited at Cliniques Universitaires Saint Luc (CUSL), Brussels, and divided into two groups: cognitively normal (CN) and AD-diagnosed groups. Cognitively normal individuals had no evidence of cognitive impairment and had a Mini-Mental State Examination (MMSE) test superior or equal to 27. The MMSE is a widely used cognitive test that is employed as routine practice at the memory clinic of CUSL. As all AD patients had previously undergone MMSE assessments, we chose to perform MMSE tests in the CN population to facilitate result comparisons between the two groups. All participants also needed to be free of neurological and psychological troubles, previous history of stroke, brain lesions, or epileptic episodes. Patients with AD were recruited based on the results of their lumbar puncture for AD biomarkers and neuropsychological evaluations. Criteria for AD diagnosis required cognitive impairment at least ine the memory domain (amnestic MCI or dementia) and an AD pattern on CSF biomarker analysis with: Aβ42 below 437 pg/ml, p-tau 181 above 61 pg/ml and total-tau above 381 pg/ml[17]. Discordant AD cases (normal Aβ42 or normal tau) were excluded. This analysis was performed during the routine medical checkup. All participants with AD underwent another MMSE evaluation for cognitive status at the time of blood collection. The lower age limit for both groups was 50 years old.

### 2.2. Blood sampling and plasma preparation

A standard blood test procedure was performed. Blood was collected with a 21-gauge needle and transferred to EDTA polypropylene K2 tubes (Vacuette, #455045). Immediately after collection, the tube was placed on ice and plasma isolation was performed within 2h. Blood was centrifugated at 2000g for 10 min at 4°C, and plasma was aliquoted by 500µl in cryotubes and stored at –80°C until further analysis.

### 2.3. Apolipoprotien E (APOE) genotyping

DNA genotyping was performed at the VIB-UAntwerp Center for Molecular Neurology (UAntwerp, Belgium) on blood samples. Participants were monitored for *APOE* single-nucleotides polymorphisms (SNPs) rs429358 ([C/T] substitution on chromosome 19q13.32 of the sequence GCTGGGCGCGGACATGGAGGACGTG [C/T]GCGGCCGCCTGGTGCAGTACCGCGG), and rs7412 ([C/T] substitution of the sequence CCGCGATGCCGATGACCTGCAGAAG [C/T]GCCTGGCAGTGTACCAGGCCGGGGC). Based on the two SNPs, one of the three APOE alleles (ε2, ε3, or ε4) was assigned.

### 2.4. Soluble Aβ and tau quantification in plasma

Quantification of soluble Aβ40, Aβ42 and total-tau was performed using the Neurology Plex A kit from Quanterix(R) (Neurology 3 Plex A, #101995). Each plasma sample was thawed at room temperature for 1h before SIMOA analysis in strict accordance with the manufacturer’s protocol. All assay runs were carried out with same duration by the same experimenter.

### 2.5. EVs isolation form plasma samples

We used size exclusion chromatography (SEC) qEVs columns from Izon Science Limited © to isolate EVs from blood plasma (#ICO–70). As it is critical to remove any aggregated or macro-protein contaminants associated with the EVs, EVs were purified using SEC rather than classical ultracentrifugation procedures[18]. Aliquots of plasma were placed at RT for 30min prior to EVs isolation to allow plasma to thaw completely. The columns were placed at RT for 30min before isolation. Columns were washed once with 20ml of DPBS (ThermoFisher scientific, #14190250) before loading with 500µl of plasma. DPBS (500µl) was used to elute and collect fractions. Fractions 7 to 12 were considered as EVs-containing fractions (see below for EVs characterization). The final volume of the EVs fraction collected was 3ml. 6ml of DPBS were added to eliminate the protein fractions. The column was subsequently washed with 1 ml of DPBS NaOH 0.5M – 16 ml DPBS 1x Triton 0.1% – 12 ml DPBS 1x – 2 ml of DPBS NaNa3 (0.05%), and finally stored at 4°C for further use. Isolated EVs were stored at 4°C for maximum 24h.

### 2.6. EVs characterization

Nanoparticle tracking, DELFIA-ELISA Europium, FACS and Dot Blot analyses were performed to characterize the isolated EVs. All validation steps were performed according to MISEV2018 guidelines [19] in n=3.Nanoparticle tracking analysis was used to determine the size distribution and concentration of isolated EVs. All samples were diluted 50 times in DPBS prior to analysis with the Particle Metrix ZetaView® analyzer.

#### 2.6.1. DELFIA-ELISA Europium

Further characterization was achieved by DELFIA-ELISA Europium sandwich assays with three different EV markers as inclusion markers (CD81, CD9, and CD63) and human serum albumin (HSA) as an exclusion marker. ELISA was performed on freshly isolated EVs (same day). All EVs samples were plated in a 96-well plate (Greiner, #762071) at 100µl per well and incubated overnight at 4°C. The plate was then washed 3 times with 300µl of 1x DELFIA wash buffer (PerkinElmer, #4010–0010), blocked with PBS BSA 1% for 1h30 with gentle agitation and washed three times again with DELFIA wash buffer. Primary antibodies were added: anti-CD81 (BioLegend, TAPA–1, #349502), anti-CD63 (BioRad, # MCA2142), anti-CD9 (R&D, # MAB1880), and anti-HSA (R&D, MAB1455) in PBS BSA 0.1% for 2h with gentle agitation. Plates were washed three times with DELFIA wash buffer and incubated with the secondary antibody (anti-mouse IgG1 biotin, PerkinElmer, #NEF823001EA) for 1h with gentle agitation. After three washes, streptavidin-europium conjugate (PerkinElmer, #1244–360) diluted 1/2500 in assay buffer (PerkinElmer, #1244–111) was added for 45 min, and then washed again six times. 100µl of enhancement solution (PerkinElmer, #1244–105) was added per well before reading the plate (Victor, Perkin Elmer).

#### 2.6.2. Identification of the NDEVs population by FACS analysis

The identification of the NDEVs population was done using CD56 (Neuronal cell adhesion molecule 1 or NCAM1) as a CNS marker. 500µl of plasma samples were incubated prior to EVs isolation with 6µl of CFSE-FITC (Life technologies ThermoFisher, #C34554A) for 1h30 at RT. EVs were isolated following the protocol described in section 2.5. After isolation, 200µl of EVs suspension was incubated overnight at 4°C with either anti-CD9-APC (R&D Biotech, #FAB1880R–100UG), anti-CD56-eFluor450 (ThermoFisher scientific, #48–0566–42) or both at the same time. FACS analysis was performed the next morning on NovoCyte Quanteon® flow cytometer. EVs stained with CFSE-FITC were gated based on the scatter scale and FITC signal. EV samples stained for CD9 were analyzed then to refine selection of the vesicle population, and finally double stained EVs (for CD9 and CD56) were considered as the NDEVs subpopulation.

#### 2.6.3. Dot Blot

Dot Blot was used to detect the presence of human IgG and ApoB (exclusion markers) in EV fractions after SEC. 25µl of each fraction were blotted onto 0.45µm nitrocellulose membranes and incubated for 15min at RT. Membranes were washed with TBS-Tween 0.1%, and incubated with guanidine chloride 6M for 5min at RT. After 3 more washes with TBS-T 0.5%, the membranes were blocked with TBS-0.5% BSA for 30min and incubated at 4°C overnight with primary antibodies (anti-human IgG-HRP (Duko #P0214) or anti-ApoB (Santa Cruz, # SC–13538)), at a dilution of 1/500 in TBS-BSA 5%. Membranes were washed twice for 15 min with TBS-Tween 0.5% under gentle agitation, incubated with secondary antibody (anti-mouse HRP, dilution 1/10.000 in TBS-BSA 5%) for 1h at RT and washed twice 15min with TBS-Tween 0.5% prior to detection using the Supersignal West Femto 10% – ECL 90% solution. Image acquisition was done with Fusion Solo Western Blot & Chemi Imaging (Vilber®).

### 2.7. Isolation and content analysis of CD56 positives EVs population

EVs isolated by SEC were concentrated by ultrafiltration (Pierce Protein Concentrators PES 10K, 2-6ml, (ThermoFisher scientific, #88527) at 4000g for 15min at 4°C. Concentrated EVs were resuspended in DPBS to reach a final volume of 500µl. The isolation of an EV subpopulation was carried out on 100µl of concentrated EVs. CD56 (neuronal cell adhesion molecule 1, NCAM1) positive EVs were isolated with the Exo-Flow kit (System Biosciences, #CSFLOWBASICA–1) following the manufacturer’s protocol, with biotinylated anti-CD56 antibody (BioLegend, #318319). After elution, EVs were concentrated by centrifugation at 100.000g for 1h30 at 4°C. EV pellets were resuspended in 25µl of RIPA lysis buffer, sonicated for 1min on ice, and centrifuged (10.000g, 10 min, 4°C). The supernatant was stored at minus 80°C for further analysis. Concentrations of Aβ42, Aβ40 and total-tau were determined using SIMOA (Neurology 3 plex A kit). In order to compare the results, we had to set a protocol indicating that the same quantity of EVs was isolated each time when using the Exo-Flow kit. To that end, we isolated as described above EVs expressing CD56 (NDEVs) by Exo-Flow. We stained EVs with CFSE-FITC (Life technologies ThermoFisher, #C34554A) by incubating 6µl of CFSE-FITC with 500µl of plasma for 1h30 on a rocking wheel. Six different quantities of EVs were used to set the number of EVs giving signal saturation, for which the same and maximal number of EVs is considered to be linked to the beads. Using the concentrated EV fraction (500µl) obtained after ultrafiltration from a control participant, we tested respectively 100µl (± 2.10^5^ EVs), 50µl (± 1.10^5^ EVs), 10µl (± 2.10^4^ EVs), 5µl (± 1.10^4^ EVs), 1µl (± 2.10^3^ EVs), and a negative control without EVs (only beads and antibody). Nanoparticle tracking analysis (NTA) (ZetaView ®, Particle Metrix) measurement was performed to calculate the EVs number in each condition. 40µl of magnetic beads coupled to biotinylated anti-CD56 antibody were used for every condition. After isolation, bead-EVs complexes were analyzed by FACS (BD FACS Canto II®) to evaluate the FITC signal (associated to EVs signal). Results are presented in supplemental data (see Supplementary Fig. 1). Bead saturation occurred at 100µl of EVs. Our measurements indicated that using 100µl of EVs after SEC and concentration to 500µl total volume was a reliable method for obtaining a consistent and maximal number of NDEVs under our experimental conditions.

### 2.8. Statistical analysis

Statistical analyses were performed using GraphPad Prism 10 software. Parametric tests were performed when data followed a normal distribution. Otherwise, non-parametric tests were performed. When two groups were compared, parametric Student’s t-test or non-parametric Mann Whitney test were used. For correlations analyses, parametric Pearson’s test and non-parametric Spearman’s test were used. Simple linear regression was used to illustrate the correlations on the graphs. Significance is indicated as: ns = non-significant, \**P* < .05, \*\**P* < .01, \*\*\**P* < .001 and \*\*\*\**P* < .0001.

## 3. Results

We first collected plasma samples from 236 CN participants and 55 AD patients and established an accurate and reliable protocol to isolate EVs from those samples, in order to discriminate free-circulating soluble AD biomarkers and those present in blood vesicles originating from the central nervous system. Descriptive parameters of all different groups are listed in Table 1, distinguishing among CN individuals *APOE* e4 carriers (n=68) and *APOE* e4 non-carriers (n=168). The full characterization of EVs isolated from CN is shown in Fig. 2.

**Table 1:** Study’s population characteristics. NOTE. Abbreviations: MMSE = Mini–Mental State Examination. Cognitive assessments were performed within 2 months of the blood draw. *APOE* e4 carriers (ε4+) and *APOE* e4 non–carriers (ε4–) indicate individuals carrying at least one e4 allele of the *APOE* gene (+) or none (–).

| Characteristics | Cognitive normal (CN, n = 236) |  | AD patients (AD, n = 55) |
| --- | --- | --- | --- |
| | *APOE* ε4- (n = 168) | *APOE* ε4+ (n = 68) | |
| Age at plasma collection<br>(years, mean ± SD) | 66.9 ± 7.5 | 65.3 ± 7.8 | 70.09 ± 8.8 |
| Female (%) | 116, 69 | 40, 59 | 30, 54.55 |
| MMSE (out of 30, mean ± SD) | 28.7 ± 1 | 28.6 ± 1 | 24.24 ± 4.33 |

### 3.1. AD soluble plasma markers vary according to APOE ε4 genotype or AD status

AD soluble plasma markers were measured across the cohort. Fig. 1 shows the quantification of soluble Aβ40, Aβ42 and total-tau in plasma. We observed a significant decrease in soluble plasma Aβ42 and Aβ42/Aβ40 ratio in CN *APOE* ε4 carriers when compared to CN *APOE ε4–* (Fig. 1.B, *P* = .019 for Aβ42 and Fig1.D *P* = .030 for Aβ42/Aβ40). No differences in soluble Aβ40 (Fig. 1.A, *P* = .89) or total-tau (Fig. 1.C, *P* = .83) were observed between CN *APOE* ε4 carriers and non-carriers. In AD patients, we observed significantly lower concentrations of soluble Aβ42 (Fig. 1.F; *P* = .012), and higher concentrations of total-tau (Fig. 1.G*, P* = .005) than in CN (*APOE* carriers and non-carriers pooled together). The Aβ42/Aβ40 ratio (Fig. 1.H) was significantly decreased in AD patients when compared to CN (*P* < .0001). Of note, the concentration of soluble plasma Aβ40 was also higher in AD patient (Fig. 1.E, *P* < .001). Values distributions of each biomarker quantified are represented in Fig. 1.E’ to H’. These two Gaussian distribution models illustrate the fold change in the mean values. The most robust fold change (1.23) appears for the soluble Aβ42/Aβ40 ratio which appears as a prime candidate for clinical diagnostic applications.

**Figure 1:**
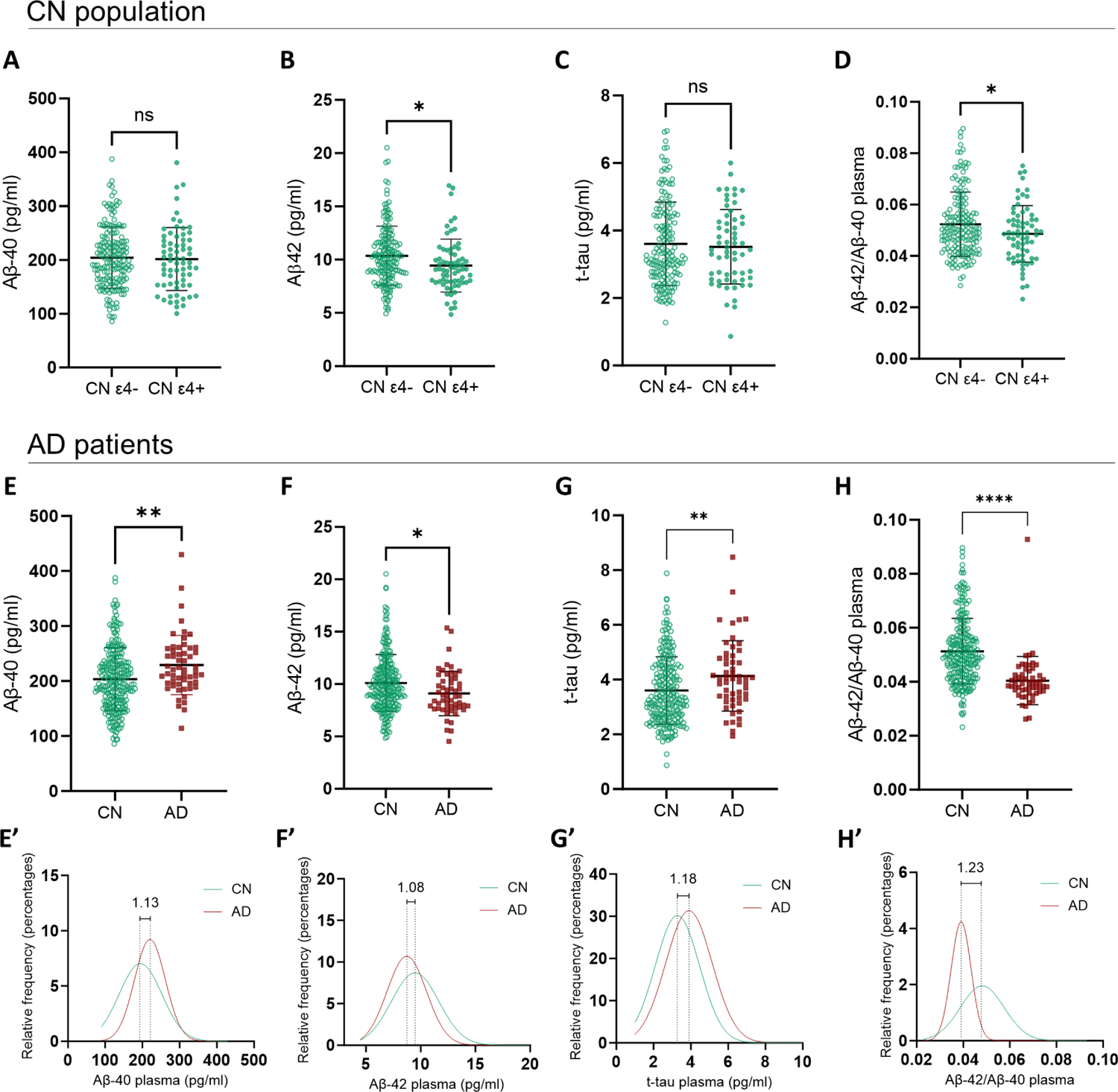
Quantification of soluble amyloid β (Aβ) 42, Aβ40 and total–tau circulating in plasma by SIMOA (Neurology 3 plex A, Quanterix). NOTE. **A** to **D:** comparison of soluble plasma AD biomarkers in CN *APOE* ε4 carriers and ε4 non– carriers. **B.** Plasma soluble Aβ42 is significantly decreased in *APOE* ε4 carriers (student’s t test, *P* value = .0186); **D.** Plasma ratio Aβ42/Aβ40 is also significantly lower in CN *APOE* ε4 carriers (student’s t test, *P* value = .0299), **A** and **C.** there were no significant differences regarding plasma soluble Aβ40 and total–tau. **E** to **H:** comparison of soluble plasma AD biomarkers in AD and CN participants. Gaussian distribution of the values with fold change of the mean value is presented under each graph (**E’ to H’**). **E.** Plasma soluble Aβ40 is significantly higher in AD patients (student’s t–test, *P* value = .0027), **F.** Aβ42 is lower in AD patients (student’s t–test, *P* value = .012), **G.** total–tau is higher in AD patients (student’s t–test, *P* value = .0052); and **H.** plasma ratio Aβ42/β40 is significantly reduced in AD patients when compared to CN participants (student’s t–test, *P* value < 0.0001).

**Figure 2:**
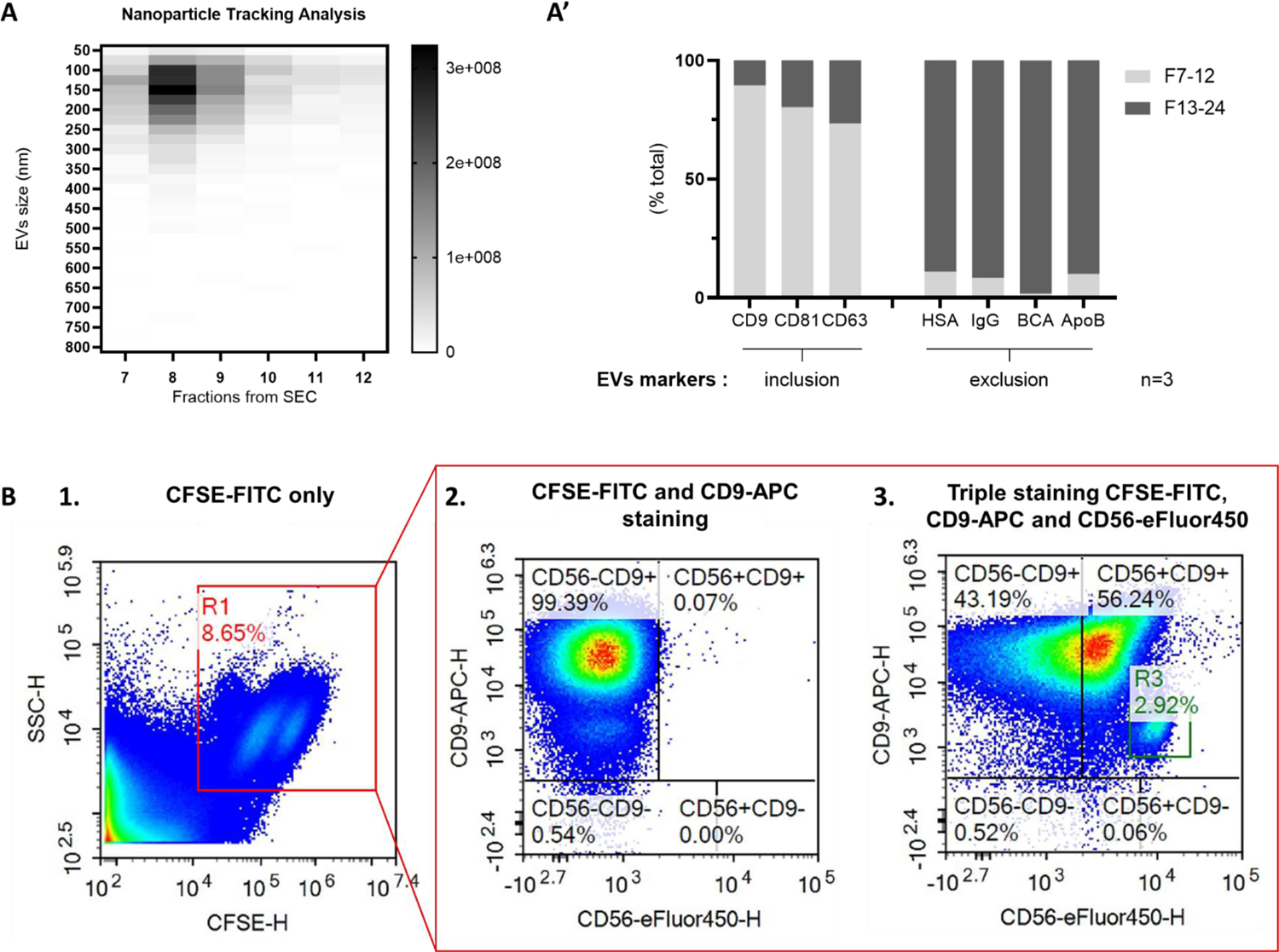
Extracellular vesicles characterization. NOTE. **A.** Nanoparticle tracking analysis (NTA) after size exclusion chromatography of 500µl of plasma. Nanoparticles were only detected in elution fractions 7 to 12 (only these fractions are displayed on the graph here). NTA provided the number and size of particles detected. The mean size of the particles was 150nm, that corresponds to the expected size of extracellular vesicles (EVs). **A’.** The particles were further characterized by using EVs inclusion markers (CD9, CD81 and CD63) and exclusion markers measured by DELFIA–ELISA (HAS) or human IgG and ApoB measured by dot blot. Protein content was measured by bicinchoninic acid assay. EVs inclusion markers were highly present in fractions 7 to 12 whereas exclusion markers were almost absent. **B.** Characterization of the neurally– derived EVs (NDEVs) population by flow cytometry (NovoCyte Quanteon). **B.1.** Full EVs population scatter plot using CFSE–FITC staining (x–axis) and side scatter measurement (SSC, y–axis). Gating (R1) was performed to isolate EVs (FITC–positive). **B.2.** Double staining was performed using CFSE– FITC and CD9–APC, as validation step for the EVs population scattering. 99% of the events present in the R1 gate (positive for CFSE–FITC) were positive for CD9–APC, which confirms that the events detected in R1 are EVs. **B.3.** Triple staining for NDEVs population detection was performed using CFSE–FITC, CD9–APC and CD56–eFluor450. The estimated NDEVs population (R3) represent 3% of the total EVs population (R1).

### 3.2. NDEVs represent 3% of the EVs circulating in plasma

To validate our method of EV purification, we first characterized the EVs isolated from the plasma of CN participants. Particle concentration and size distribution was determined in each SEC fraction with Nanoparticle Tracking Analysis (Fig. 2.A). Particles were only detected in fractions 7 to 12, with higher concentrations in fractions 8 and 9. The particles detected ranged from 50 nm to 350 nm in size, with most particles measuring 150-200 nm, corresponding to the expected size of EVs. Additional experiments indicated that inclusion markers were strongly enriched in fractions 7 to 12. The expression profiles of these different EV markers were quantified using the ELISA-DELFIA immunoassay (see Methods, Section 2.7), revealing 89% of total CD9 signal, 80% of total CD81 signal, and 73% of total CD63 signal (Fig. 2.A’). The exclusion markers were predominantly found in fractions 13 to 24, wherein 89% of Human Serum Albumin, 92% of total IgG, 98% of total protein, and 90% of ApoB were detected. Next, we characterized the subpopulation of brain-derived EVs referred to here as Neurally-derived EVs or NDEVs using flow cytometry. EVs were stained with CFSE-FITC and selected based on the size scatter scale and FITC signal (Fig. 2.B). We confirmed that the population selected consisted of EVs by performing double staining with CFSE-FITC and CD9-APC, where 99.36% of the total EV population tested positive. Finally, we added CD56-eFluor450 staining to the CFSE-CD9 condition to quantify the number of EVs expressing Neuronal Cell Adhesion Molecule 1 (NCAM1 or CD56) and thus derived from the CNS. Only 3% of the CFSE-CD9 positive population tested positive for CD56 (NCAM1), indicating that the NDEVs population represented around 3% (1/30) of the EVs circulating in the bloodstream (Fig. 2.B.3). When only stained with CFSE-FITC and CD56-eFluor450 (not CFSE-CD9, data not shown), up to 4% of CFSE-FITC positive events were positive for CD56-eFluor450. This indicates the presence of NDEVs in plasma, representing between 3% and 4% of the total EVs circulating in plasma.

### 3.3. Relative quantification of NDEVs in plasma

We next investigated whether the number of NDEVs could be used as an AD biomarker in plasma samples. Relative plasma NDEV levels were measured in a subpopulation of CN participants (n = 62, mean age of 64.9 [7.02], mean MMSE of 28.7 [1.15], 32 women [60.4%], *APOE* ε4+/ε4– [27/35]) and AD patients (n = 54, mean age of 70.1 [8.82], mean MMSE of 24.2 [4.3], 30 women [54.5]). Results are shown in Fig. 3. No significant differences were observed in CN individuals regarding the *APOE* ε4 genotype (Fig. 3.A; *P* = 0.93). Importantly, there was no correlation between age and plasma NDEVs levels in CN *APOE* ε4 carriers or non-carriers (Fig. 3.B). Plasma NDEVs levels were not significantly different in AD patients compared to CN individuals (Fig. 3.C, *P* = 0.28). In agreement with this observation, we did not observe any correlation between NDEVs and MMSE scores in the AD group (Fig. 3.D).

**Figure 3:**
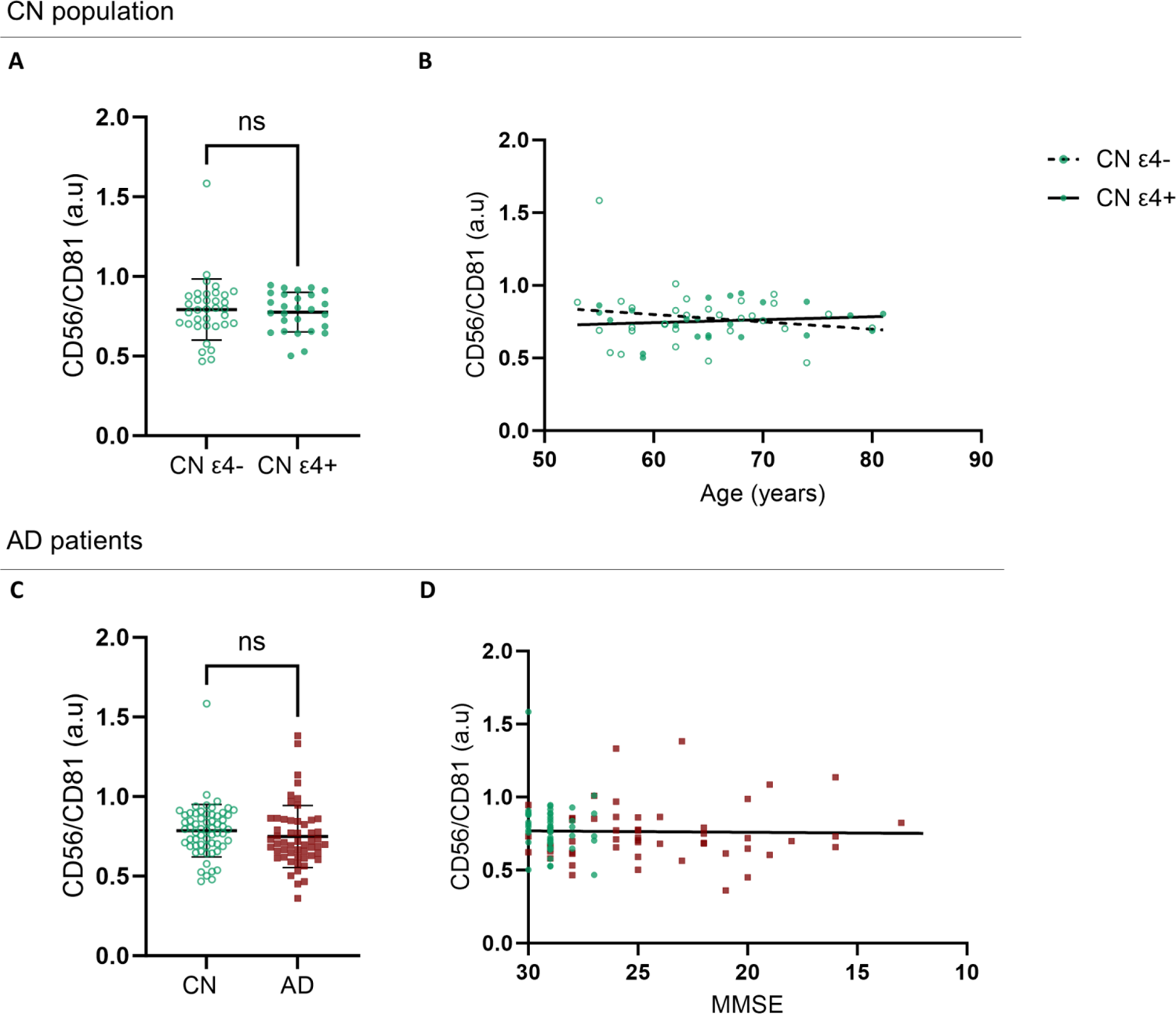
Plasma NDEVs quantifications measured by ELISA–DELFIA immunoassay. NOTE. The relative numbers of NDEVs is calculated as the ratio of CD56 signal on CD81 signal**. A.** Measurement of NDEVs in plasma from CN *APOE* ε4 carriers and non-carriers (Mann–Whitney test, P value = 0.93); **B.** Age-related variation of plasma NDEVs quantity from CN *APOE* ε4 carriers and non-carriers (simple linear regression, Spearman correlation test, r = –0.006 and *P* value = .98 for ε4 non– carriers; r = 0.13 and *P* value = .55 for ε4 carriers). **C.** Measurement of NDEVs in plasma from CN participants and AD patients (student’s t–test, *P* value = .28); **D.** Variation of plasma NDEVs quantity with cognitive loss progression (CN participants are displayed in green, and AD patients in red, Simple linear regression with Spearman correlation test, r = 0.02, *P* value = .84).

### 3.4. Inverse correlation between soluble amyloid biomarkers and NDEVs level in CN individuals

Our results indicated that plasma NDEVs numbers do not significantly change (i) with age in CN individuals, (ii) between CN individuals and AD patients, (iii) with decreased MMSE performance. This indicates that the number of plasma NDEVs do not appear as a clinical biomarker to discriminate between CN individuals and AD patients or to monitor cognitive decline [20]. Considering that EVs and in particular exosomes can be produced in multivesicular bodies (MVBs) formed in early endosomes [21] where APP amyloidogenic processing occurs [22], recent findings showed that Aβ is readily present in small extracellular vesicles [23]. We next investigated the correlation between AD soluble plasma biomarkers, NDEVs number and their content in AD biomarkers (i.e. Aβ42, Aβ40 and total tau). Results are summarized in Fig. 4 and detailed scatter plots can be found in Supplementary Fig. 2. In CN participants (displayed in green on Fig. 4), we found a significant negative correlation between the number of plasma NDEVs and the concentration of soluble plasma Aβ42 and Aβ40 (Fig. 4, A. and B., Pearson’s correlation; for Aβ42: *P* < .0001, r = –0.51; for Aβ40: *P* = .0003, r = –0.45). For Aβ42, up to 27% of the variation observed among CN participants was explained by the number of NDEVs circulating in the blood (simple linear regression, R² = 0.27); for Aβ40, it was close to 20% (simple linear regression, R² = 0.20). In parallel, Aβ content was measured in NDEVs (Fig. 4, A’. and B’.). Experimental procedures were set to ensure that equal numbers of EVs were measured in each condition. Under these conditions, high numbers of NDEVs were correlated with high individual content in Aβ42 and Aβ40 (Spearman’s correlation, for Aβ42: *P* = .015, r = 0.47, for Aβ40: *P* = .007, r = 0.52). In AD patients (displayed in red on Fig. 4), soluble Aβ42 and Aβ40 concentrations in plasma were not correlated to the quantity of plasma NDEVs (Fig. 4, A. and B.) (Pearson correlation, for Aβ42: *P* = .22, r = –0.17, for Aβ40: *P* = .26, r = –0.15). However, we saw a strong correlation between NDEVs quantity and their content in Aβ in AD patients (Sperman’s correlation, for Aβ42: *P* = .0065, r = 0.69; and for Aβ40: *P* = .0082, r = 0.72).

**Figure 4:**
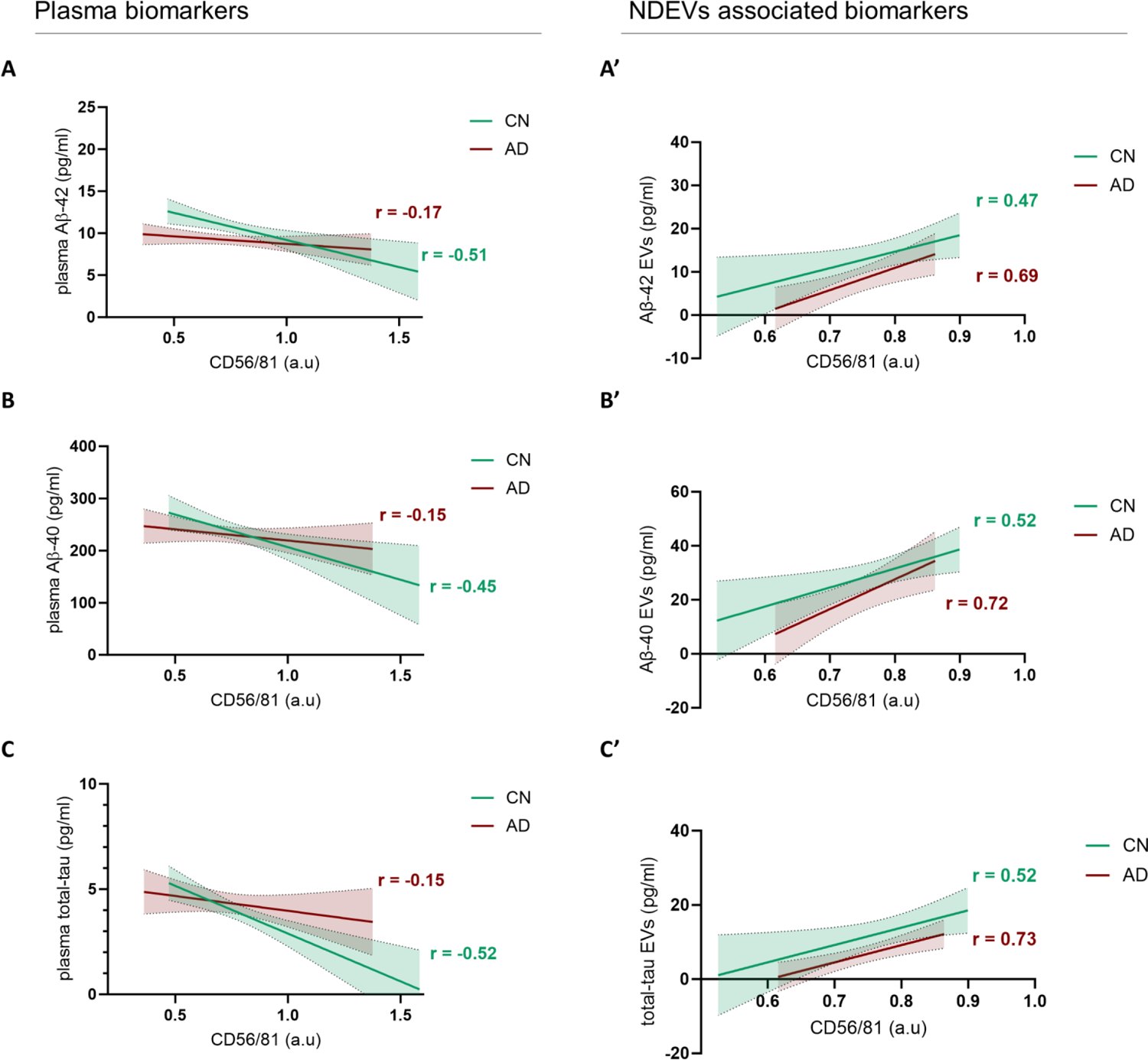
Correlations between plasma biomarkers, plasma NDEVs level and NDEVs associated biomarkers. NOTE. **Left column (Plasma biomarkers):** correlations between plasma soluble AD biomarkers quantity and plasma relative NDEVs quantity (ELISA signal ratio CD56/81) in AD (red) and CN (green) populations. Statistics: Pearson correlation with simple linear regression model (95% confidence intervals displayed for each curve). **A.** Strong correlation in CN for soluble Aβ42 and NDEVs quantity in plasma (*P* value = .031), loss of correlation in AD (*P* value = .22); **B.** significant correlation between Aβ40 and NDEVs quantity in plasma of CN (*P* value = .009), loss of correlation in AD (*P* value = .26); **C.** strong correlation for t–tau and NDEVs in plasma of CN (*P* value = .0002), and loss of correlation in AD (*P* value = .25). **Right column (NDEVs associated biomarkers):** correlations between NDEVs content measured by SIMOA (N3PA kit) in the same number of EVs (± 2.10^5^ EVs, cf. Appendix Figure 2 for details), and the relative NDEVs quantity in plasma (ELISA signal ratio CD56/81) in AD and CN populations. Statistics: Spearman correlation with simple linear regression (95% confidence intervals are displayed in each curve). **A’.** Strong correlation between Aβ42 carried in NDEVs and NDEVs relative quantity in plasma for both CN (*P* value = .031) and AD (*P* value = .0048); **B’.** strong correlation between Aβ40 carried in NDEVs and NDEVs relative quantity in plasma for both CN (*P* value = .014) and AD (*P* value = .0065); and **C’.** strong correlation between t–tau carried in NDEVs and NDEVs relative quantity in plasma for both CN (*P* value = .027) and AD (*P* value = .002).

### 3.5. Inverse correlation between soluble total-tau and NDEVs level in CN volunteers

As for Aβ, soluble plasma total-tau in CN individuals negatively correlated with the number of plasma NDEVS (Fig. 4.C, Pearson’s correlation, *P* < .0001, r = –0.52). Measuring EVs content for total-tau, we found a significant correlation between the quantity of total-tau in NDEVs and the number of plasma NDEVs (Fig. 4.C’, Pearson’s correlation, *P* < .05, r = 0.49) as observed for Aβ42 and Aβ40. In AD patients, there was no significant correlation between NDEVs levels and soluble plasma total-tau (Fig. 4.C), but the content of total-tau in NDEVs strongly correlated with NDEVs quantity (Fig. 4.C’, Spearman’s correlation, *P* < .0001, r = 0.73).

### 3.6. No differences of soluble AD biomarkers quantity linked to NDEVs between AD and CN individuals

We further investigated the potential use of NDEVs-associated amyloid and tau as AD biomarkers. Aβ42, Aβ40 and total-tau quantifications were done on the same number of NDEVs for each participant (see Supplementary Fig. 1 for details). For the three biomarkers tested, concentrations were not significantly different in AD compared to CN participants (Fig. 5. A, B and D) (Mann Whitney test). Aβ42/Aβ40 ratio was also similar (Fig. 5. C) (Mann Whitney test) but we observed a non-significant trend for a decreased Aβ42/40 ratio in the NDEVs of AD patients.

**Figure 5:**
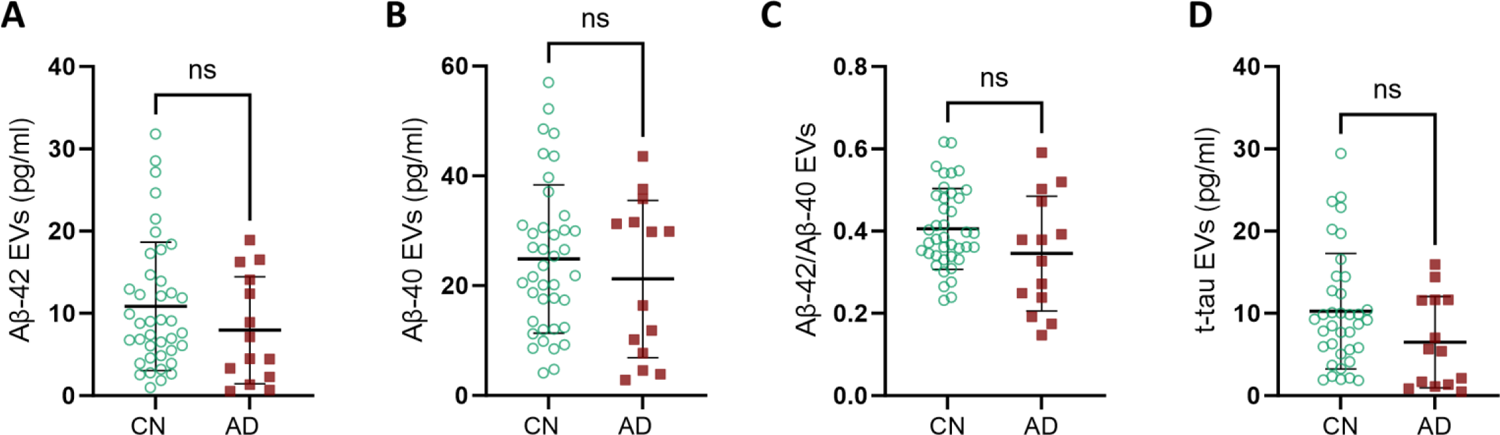
Comparison of the content in AD biomarkers of plasma NDEVs analyzed by SIMOA (N3PA kit). NOTE. For the analysis of NDEVs content, we focused on subgroups distributed across the plasma NDEVs spectrum, ranging from low circulating participants to high circulating participants. In the CN group, n=27, with a mean MMSE of 28.76 [1.27], mean age of 61.78 [14.8], 16 women [59.3%], and mean CD56/81 of 0.768 [0.10]. In the AD group, n=14, with a mean MMSE of 21.9 [5.09], mean age of 66.4 [16.7], 6 women [42.8%], and mean CD56/81 of 0.742 [0.09]. Comparisons were done with Mann Whitney test. **A.** Aβ42 contained in NDEVs do not change in AD (*P* value = .21); **B.** Aβ40 either (*P* value = .46); **C.** the ratio Aβ42/Aβ40 do not change in AD (*P* value = .14); and **D.** total–tau either (*P* value = 0.06).

## 4. Discussion

Blood biomarkers are of growing interest for early detection of AD, monitoring its progression, and measuring the effects of disease-modifying therapies. However, their implementation in clinical practice faces challenges due to low concentrations of AD biomarkers (Aβ, tau) in blood, requiring reliable measurement techniques and standardized sample preparation methods. In our study, we established a standardized protocol to measure free-circulating soluble Aβ42/40/total-tau, NDEVs, and their content in Aβ42/40/tau by SIMOA in human plasma samples, with the goal of assessing their potential as AD biomarkers. Our major findings are that (i) Aβ42/40 ratio significantly discriminates between CN individuals and AD patients; (ii) NDEVs are present in the blood (around 3% of total blood EVs), but their number does not change with age or with AD dementia; (iii) in CN individuals, there is an inverse correlation between soluble AD biomarkers and the number of NDEVs. Strikingly, the content of AD biomarkers in NDEVs does not significantly change between CN individuals and AD patients and it does not therefore appear as a diagnostic tool to identify AD patients when quantified by SIMOA.

### 4.1. Plasmatic soluble AD biomarkers

Previous studies have identified significant differences in plasma levels of Aβ40, Aβ42, and total-tau between AD patients and CN individuals[20, 24, 25]. Interestingly, we observed a slight but significant decrease in Aβ42 concentration and Aβ42/40 ratio in CN *APOE* ε4 carriers, emphasizing the connection between *APOE* ε4 and amyloid pathology. This suggests plasma Aβ42/40 ratio as a candidate biomarker to detect preclinical stages of at-risk individuals (i.e. *APOE* ε4 carriers) to develop AD. The fold change in the Aβ42/40 ratio between CN and AD patients measured in plasma (1.23) is markedly lower than the one measured in CSF[26]. Amyloid and tau concentrations in plasma are approximately tenfold lower than those in CSF[27, 28]. Conversely, the total protein content in plasma is tenfold higher than in CSF[29], rendering the quantification of plasma biomarkers more exposed to biases likely assay cross reactivities or buffering due to the binding to serum albumin present at high concentration in plasma.

Although we observed a significant increase in soluble total-tau in AD plasma samples, we found it to be a less robust biomarker than Aβ42/40 ratio. These results are consistent with many studies that support that total tau[30–32] is more efficient when used in combination with other biomarkers, such as soluble Aβ42, to identify patients with AD. Though increase in CSF total-tau is used routinely in clinic for AD diagnosis, we are fully aware that phospho-tau measurements (e.g. p-Tau 181 or p-Tau 217) are more sensitive to detect AD pathology[30], they were not possible to achieve in the triplex panel available. Further experiments would be useful to complete this biomarkers collection by testing independently p-tau biomarkers.

We must assume that only one fraction of Aβ and tau present in the CSF flows to the blood stream, or that peripheral Aβ which has been shown to regulate Aβ clearance from the CNS might interfere in the measurement. In that respect, isolating NDEVs and measuring NDEVs content is of prime interest, as they might better reflect differences between AD and CN individuals than soluble, free-circulating markers.

### 4.2. NDEVs as an AD biomarker

The analyze of NDEVS was the second outcome of this study, aiming to evaluate their potential as biomarkers for neurodegenerative conditions and their impact on soluble biomarker measurements, considering their role in amyloid and tau propagation While few studies have explored NDEV quantities as AD biomarkers, Kapogiannis et al.[33] did not observe any differences in NDEVs quantity between AD patients and CN controls using L1CAM as a neuronal marker to identify NDEVs within the total EVs pool. However, the use of L1CAM has become controversial due to its presence in a soluble form in plasma, unrelated to EVs[34]. In our research, we employed a different neuronal marker, NCAM1 (neuronal cell adhesion molecule one, also referred as CD56), expressed by neuronal cells. Despite being expressed by cell types other than neurons and especially NK cells[35], NCAM1 is currently the best available tool to isolate or quantify EVs derived from the CNS in plasma. In the present study, we employed FACS analysis for NDEVs detection as a confirmatory method, as no previous study, thus enabling the validation of NDEVs isolation and the estimation of this EVs subpopulation to comprise 3% of circulating EVs in plasma. In agreement with Kapogiannis’ study using L1CAM, we did not find any significant differences in plasma NDEVs quantity between AD and CN groups in ELISA quantifications. The production of EVs in the brain - at least those than can be monitored in the blood - does not seem to be affected by disease conditions. Consequently, levels of neuronal-derived EVs appears neither as a marker of brain aging nor as an indicator of dementia onset or progression, as no changes were observed in AD patients upon deterioration of the MMSE score. If not the number, the content of NDEVs might then reflect pathological conditions.

### 4.3. NDEVs content as an AD biomarker

We found that the levels of Aβ42, 40 or total-tau associated to NDEVs were not altered in AD patients when quantified using SIMOA. The concept of NDEVs content as an AD biomarker is an emerging field with limited available literature, and there is currently no consensus on the variations observed in AD patients. Other studies using SIMOA found similar results regarding Aβ42[33], or total tau[36] carried in NDEVs. Conversely, when quantified by ELISA, significantly elevated Aβ42 levels were reported in plasma NDEVs of AD patients[37–39]. For total-tau quantified by ELISA, similar results as for Aβ42 were reported[38, 39], but other immunoassays gave inconsistent results, with no significant differences of total-tau in NDEVs of AD patients[40]. The inconsistency in results across studies might stem from variations in the techniques used for biomarker quantifications (e.g., SIMOA, ELISA kits), NDEVs isolation (e.g., the use of NCAM1 and/or LCAM1 as primary target for NDEV immunoprecipitation), and isolation of extracellular vesicles (EVs) from plasma. Still, our study here supports the hypothesis that neither the number or the content of NDEVs measured in plasma does not discriminate AD patients from CN individuals.

### 4.4. The relation between plasma NDEVs amount, their content and soluble AD biomarkers could reflect the dynamic of biomarkers production at the neuronal levels that is impaired in AD pathology

Levels of free-circulating Aβ42, Aβ40 and total-tau in plasma inversely correlated with NDEVs quantity in CN individuals, indicating that higher NDEV numbers corresponded to lower soluble amyloid and tau concentrations, including among CN individuals at low-risk (*APOE* ε4 non-carriers). However, the quantity of biomarkers linked to NDEVs was also correlated to NDEVs number (when analyzed on the same amount of NDEVs), with higher numbers of NDEVs circulating associated to higher content in AD biomarkers. Together, these results strongly suggest a link between soluble AD biomarkers and the number of NDEVs, with biomarkers circulating either freely or embedded in EVs, but with a balance between their respective concentrations only in CN individuals. In AD patients, this correlation between soluble AD biomarkers and NDEVs was lost, reflecting potential changes in Aβ production and release during AD pathogenesis. It has been found that Aβ is present in MVBs and can be released into the extracellular space through EVs in neurons. Recent studies have demonstrated the presence of APP, β- and γ-secretase in exosomes and at the endosomal level[41–43] providing further evidence of a clear link between amyloid and EVs.

Increased soluble Aβ40 found in the plasma of AD patients may reflect and overall increase in Aβ production in AD condition. Decreased soluble Aβ42 and consequently more pronounced decrease in the Aβ42/40 ratio is likely to reflect Aβ42 aggregation and deposition in AD pathology, leaving less Aβ42 available. Aβ42 aggregates are not detected in the current assays but would be very valuable tool to track AD pathology in blood samples. If plasma Aβ measurement truly reflects Aβ production and release in the brain parenchyma, our result would indicate an equilibrium existing between the release of soluble Aβ and Aβ embedded in vesicles in healthy conditions (CN). This balance seems to be broken in AD conditions, with soluble Aβ42 decrease more likely to be a consequence of amyloid-β deposition the brain than vesicular Aβ (NDEVs) that does not significantly decrease in AD condition. The possible interconnection between these observation ad underlying mechanisms is summarized in Fig 6.

**Figure 6:**
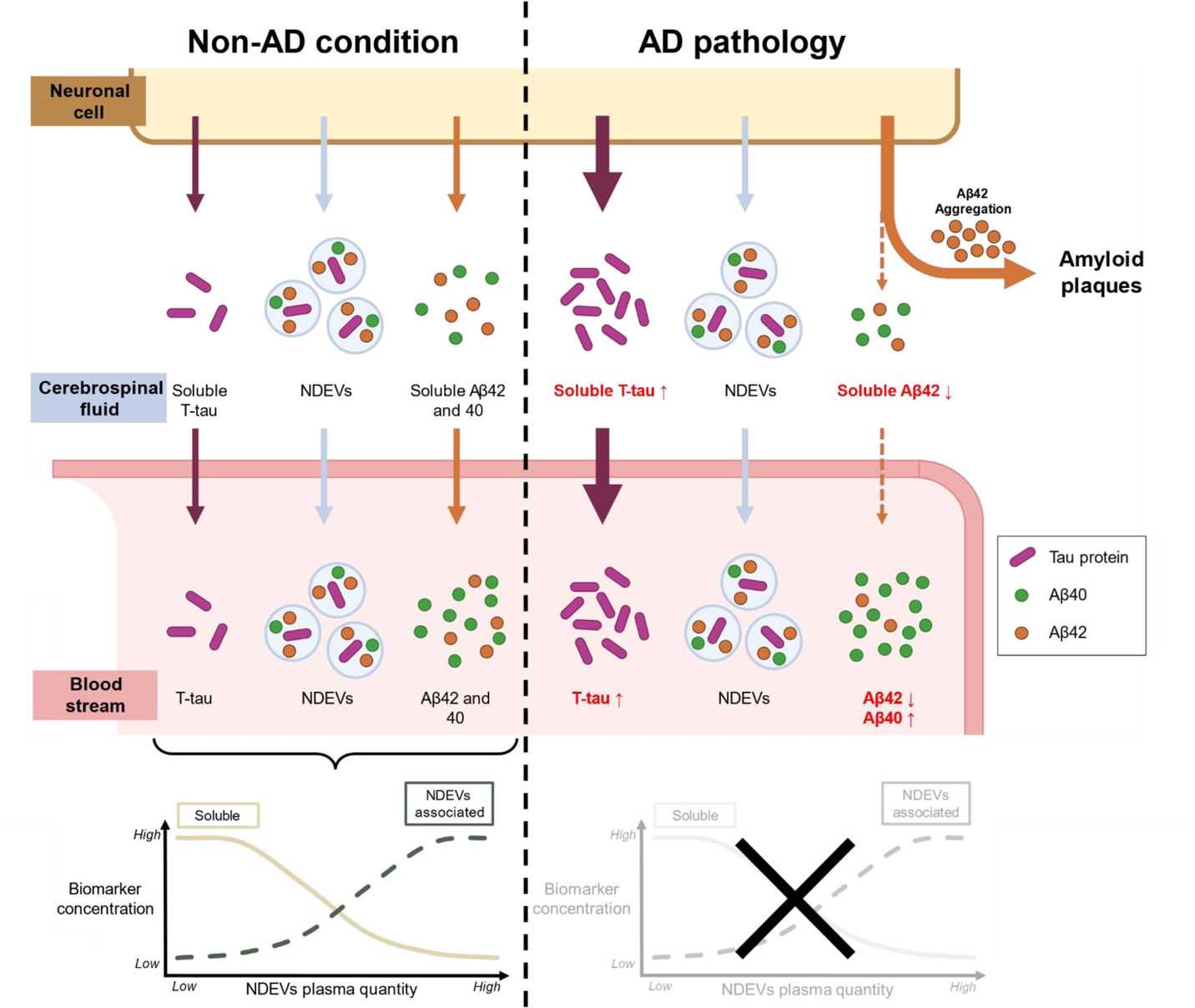
Summary of the main findings. NOTE. In the non–AD condition (**left part**), tau protein and amyloid are found either circulating in plasma as soluble (freely circulating) or linked to NDEVs. For total–tau, Aβ42 and Aβ40, the proportion of soluble plasma biomarkers is highly correlated with plasma NDEVs quantity and their content in biomarkers. This distribution between NDEVs and the soluble compartment is balanced in non–AD (as represented in the **bottom–left graph**), reflecting the mechanisms of tau and amyloid production at the neuronal level observed in in vitro models. In AD pathology (**right part**), the release of Aβ and tau is increased, leading to an increased concentration of tau, and a transitional increase of Aβ42–40, in the CSF. This amyloid is more prone to aggregation and will form amyloid plaques, resulting in a decreased quantity in the CSF. The same pattern is observed in the plasma of AD patients, but the dynamics regarding plasma NDEVs are here imbalanced by the development of AD pathology (as represented in the **bottom–right graph**). The content or number of NDEVs seems not to be good AD biomarkers, whereas variations in plasma soluble biomarkers are more accurate for AD diagnosis.

### 4.5. Confounding factors

In our study, we decided to use plasma samples instead of serum for the quantification of amyloid and total tau. Plasma is collected under anticoagulation conditions, preventing clot formation. Clotting events can lead to protein precipitation, thereby interfering with protein quantification[44]. Plasma sample also have the advantage of minimizing hemolysis in comparison to serum. Since blood cells are a potential source of amyloid, it is important to reduce external contamination for reliable amyloid quantifications in plasma[45]. Plasma samples are thus the best material to standardize measurements in blood sample and maximize their accuracy.

## 5. Conclusion

In conclusion, our study provides valuable insights into the challenges regarding the use of blood biomarkers for AD diagnosis. The intricate relationships observed between NDEVs and soluble AD biomarkers emphasize the need for comprehensive consideration when interpreting biomarker levels in CN participants.

## Data Availability

All data produced in the present study are available upon reasonable request to the authors

## Acknowledgments

We thank all participants and the Stichting Alzheimer’s Onderzoek for their help in our study. We thank Daniela Savina, Yasmine Salman, Lisa Quenon and Lara Huyghe for their technical supports.

## Supplementary Data

**Figure 1:**
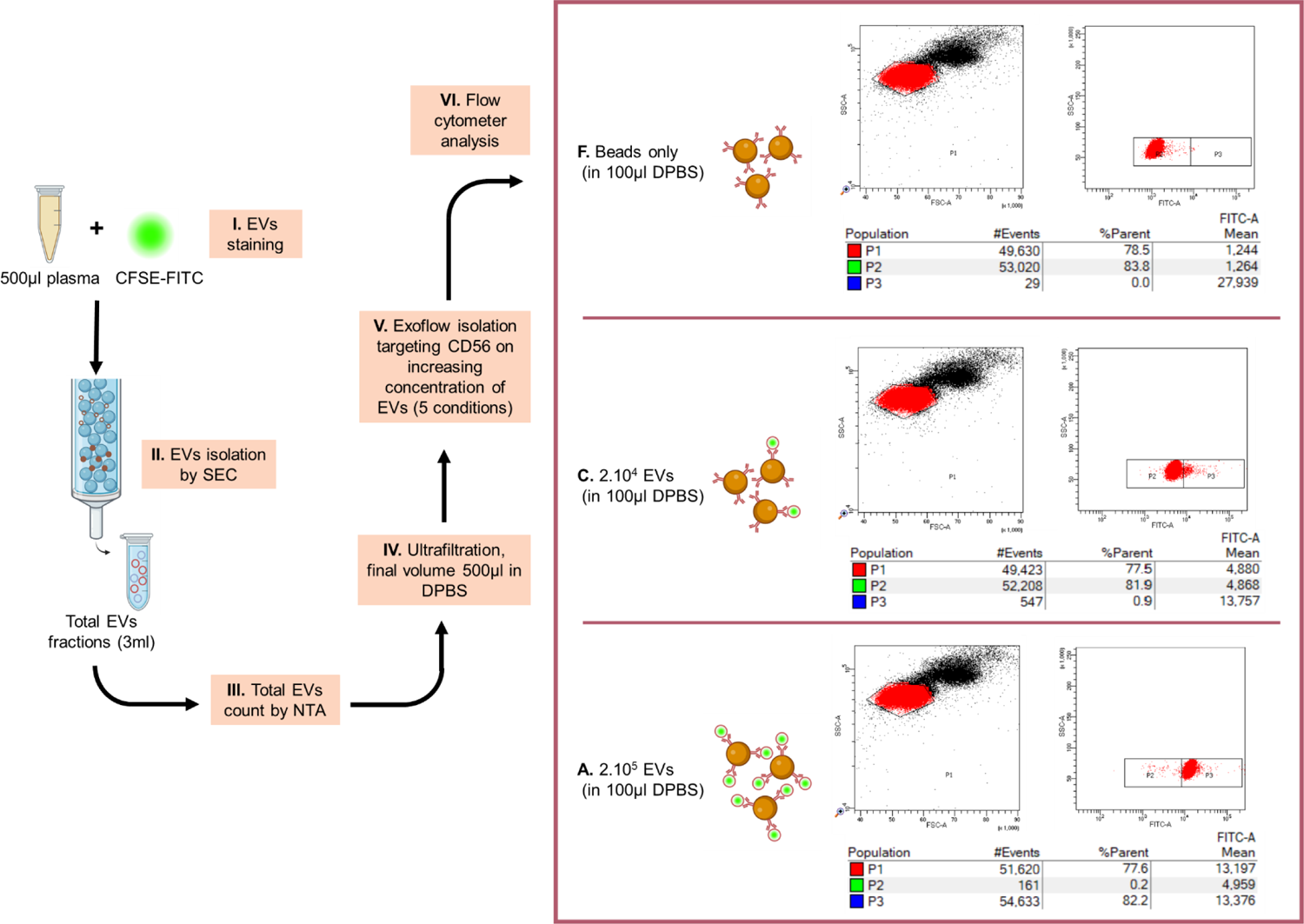
Validation protocol for NDEVs isolation using Exoflow technique. NOTE. **I.** Plasma sample was incubated with 6µl of CFSE–FITC at RT for 2h. **II**. EVs were isolated by size exclusion chromatography, by collecting EVS fractions (F7 to 12 = 3ml total volume). **III.** Number of EVs were count by NTA to determine EVs quantity to use in Exolfow protocol. **IV.** 3ml of eluted EVs were concentrated by ultrafiltration (10kDa filter, 4°C, 45min at 4000g), final volume was 500µl. **V.** Exoflow with anti–CD56 antibody was performed in 5 different conditions: A. 100µl of concentrated EVs (+/–2.10^5^ EVs), B.50µl (+/– 1.10^5^ EVs), C.10µl (+/– 2.10^4^ EVs), D.5µl (+/– 1.10^4^ EVs), E.1µl (+/–2.10^3^ EVs) and F. magnetic beads without EVs. We used the same amount of beads solution (40µl) and antibody (10µl, dilution 1/5). Exoflow isolation was performed following manufacturer protocol (System Biosciences, #CSFLOWBASICA–1). **VI.** Post isolation beads were analyzed by flow cytometry for FITC signal. Beads population was selected based on size scatter and forward scatter (population P1), and with negative FITC signal (population P2). Positive population (P3) was considered positive above log10^4^ of FITC signal. Saturation point was reach with condition A. 2.10^5^ EVs.

**Figure 2:**
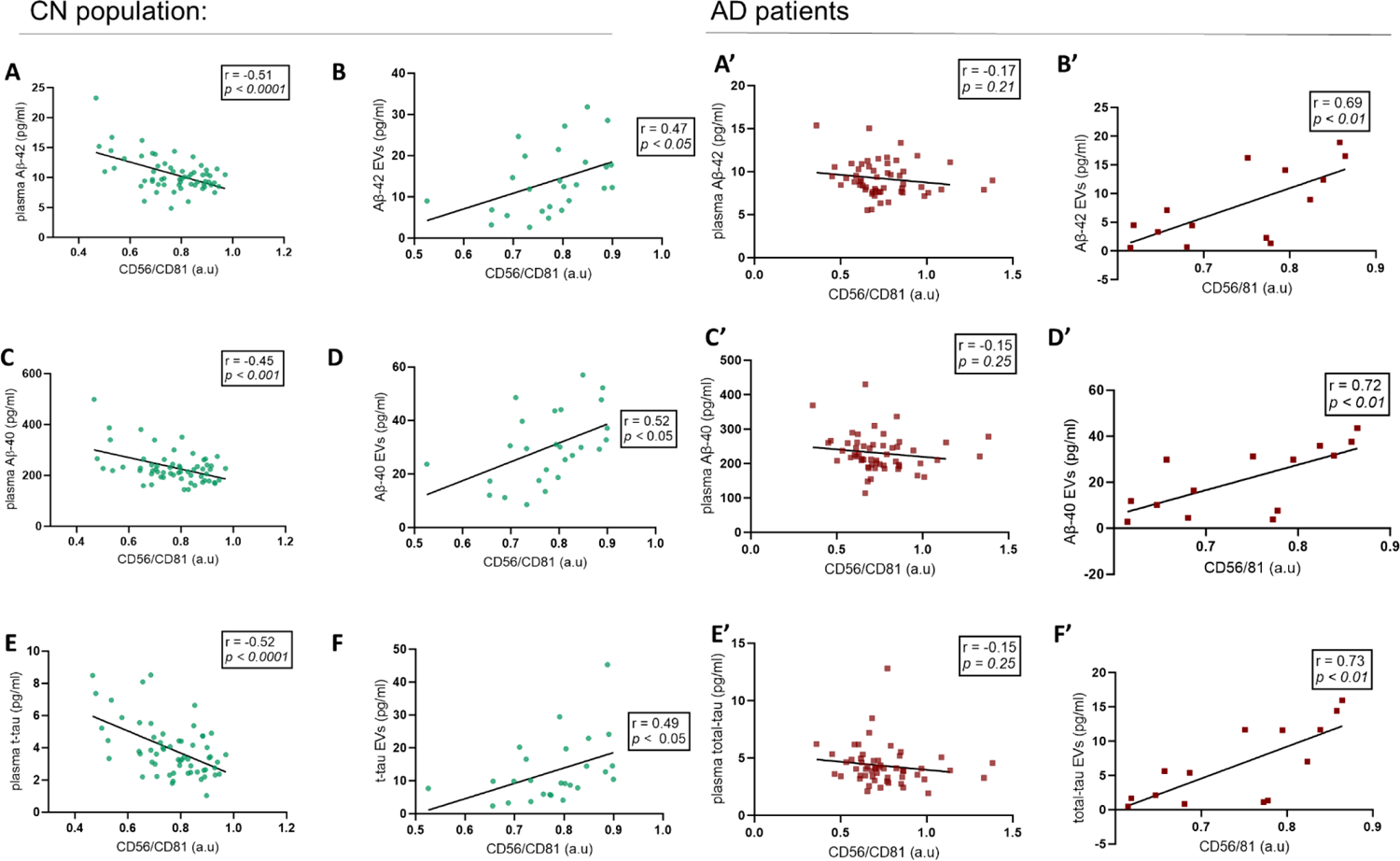
Detailed scatter plots of the correlation between plasma soluble biomarkers quantity, plasma NDEVs quantity and their content in AD biomarkers. NOTE. Population details for figures A, A’, C, C’, E and E’ can be found on paragraph 3.2. For the analysis of NDEVs content, we focused on subgroups distributed across the plasma NDEVs spectrum, ranging from low circulating participants to high circulating participants. In the CN group, n=27, with a mean MMSE [SD] of 28.76 [1.27], mean age of 61.78 [14.8], 16 women [59.3%], and mean CD56/81 of 0.768 [0.10]. In the AD group, n=14, with a mean MMSE of 21.9 [5.09], mean age of 66.4 [16.7], 6 women [42.8%], and mean CD56/81 of 0.742 [0.09].

